# COVID-19 transmission in a theme-park

**DOI:** 10.1101/2021.04.15.21255560

**Authors:** Elena Aruffo, Safia Athar, Angie Raad, Md Afsar Ali, Mohammed Althubyani, Christopher Chow, Mahmuda Ruma, Chao Liu, Jianhong Wu, Ali Asgary, Jude D. Kong, Jane M. Heffernan

**Author notes:** These authors contributed equally to this work (Elena Aruffo, Safia Athar, Angie Raad).

## Abstract

**Background:** As COVID-19 vaccination coverage increases, public health and industries are contemplating re-opening measures of public spaces, including theme-parks. To re-open, theme-parks must provide public health mitigation plans. Questions on implementation of public health mitigation strategies such as park cleaning, COVID-19 testing, and enforcement of social distancing and the wearing of personal protective equipment (PPE) in the park remain.

**Methods:** We have developed a mathematical model of COVID-19 transmission in a theme-park that considers direct human-human and indirect environment-human transmission of the virus. The model thus tracks the changing infection/disease landscape of all visitors, workers, and environmental reservoirs in a theme park setting.

**Findings:** Models results show that theme-park public health mitigation must include mechanisms that reduce virus contamination of the environment to ensure that workers and visitors are protected from COVID-19 transmission in the park. Thus, cleaning rates and mitigation of human-environment contact increases in importance.

**Conclusion:** Our findings have important practical implications in terms of public health as policy- and decision-makers are equipped with a mathematical tool that can guide theme-parks in developing public health mitigation strategies for a safe re-opening.

## Introduction

Epidemiological data on Severe acute respiratory syndrome coronavirus-2 (SARS-CoV-2) suggests that this novel virus can be transmitted both directly from human to human via respiratory droplets [22, 29], and indirectly through a contaminated environment [22, 29] (It can survive on surfaces for several hours to days ([7]). In the absence of pharmaceutical interventions, to control the rapid spread of the virus, health authorities worldwide have implemented strict social distancing measures, and have recommended hand washing, the use of PPE (i.e., masks), and the increased sanitization of public spaces [8, 17]. Many of these measures were adopted early on in the pandemic, along with the closing of non-essential businesses [35, 34, 30, 33, 39, 5]. However, as local and global economies have been suffering, various countries have eased lock-down measures, allowing non-essential businesses to resume their services [35, 34, 30, 33, 39, 5] while adhering to specific public health guidelines. Stricter measures were implemented after new variants were discovered in Fall 2020 [2, 42, 10]. According to recent studies [40,20, 12, 28, 41], the new variants are more contagious than the one known so far, with an increase of transmissibility ranging between 35% and 71% [12, 28, 41].

Amusement parks are among the facilities that have been allowed to reopen in various jurisdictions, and others are following suit over time. Given the expected large number of visitors to the parks, it is necessary to implement safety measures to mitigate the spread of the virus. The following mitigation strategies have been put in place in various theme-parks that have reopened: limiting the number of guests, increasing cleaning and sanitization services, introducing a mandatory mask mandate for visitors, mandating the use of other personal protective equipment (PPE) for workers, temperature checks for visitors and workers upon gate entry, and regular hand sanitization routines [31, 32, 16, 27, 19, 21].

Mathematical models of SARS-CoV-2 direct transmission have highlighted the importance of adopting social distancing measures to reduce human-to-human contact ([37,26]). Considering an indirect mode of transmission only, Xie et al. ([43]), examined the importance of adopting frequent sanitization routines to reduce transmission via contaminated surfaces ([43]). Using a hybrid multi-scale model including direct and indirect transmission, Bounchinta et al. ([4]) showed that infected surfaces can play a crucial role in disease transmission. We extend the study of direct and indirect transmission to a theme-park setting where both transmission routes will affect COVID-19 spread. Here within, we quantify the effects of environmental sanitization in reducing indirect transmission, and the effects of social distancing and personal protective equipment (PPE) adherence rates in reducing direct transmission of the virus. We also assess the efficacy of COVID-19 symptom checking (i.e., temperature checks) at the park gate. A multi-patch model is employed to capture transmission outcomes in two different areas of the park. Both areas include a queue and a ride environment.

## Methods

### Model formulation and assumptions

A model flow diagram is presented in Fig. 1 (see Figure S1 for a more detailed diagram). A detailed description of the mathematical model is provided in the Appendix.

**Fig. 1.**
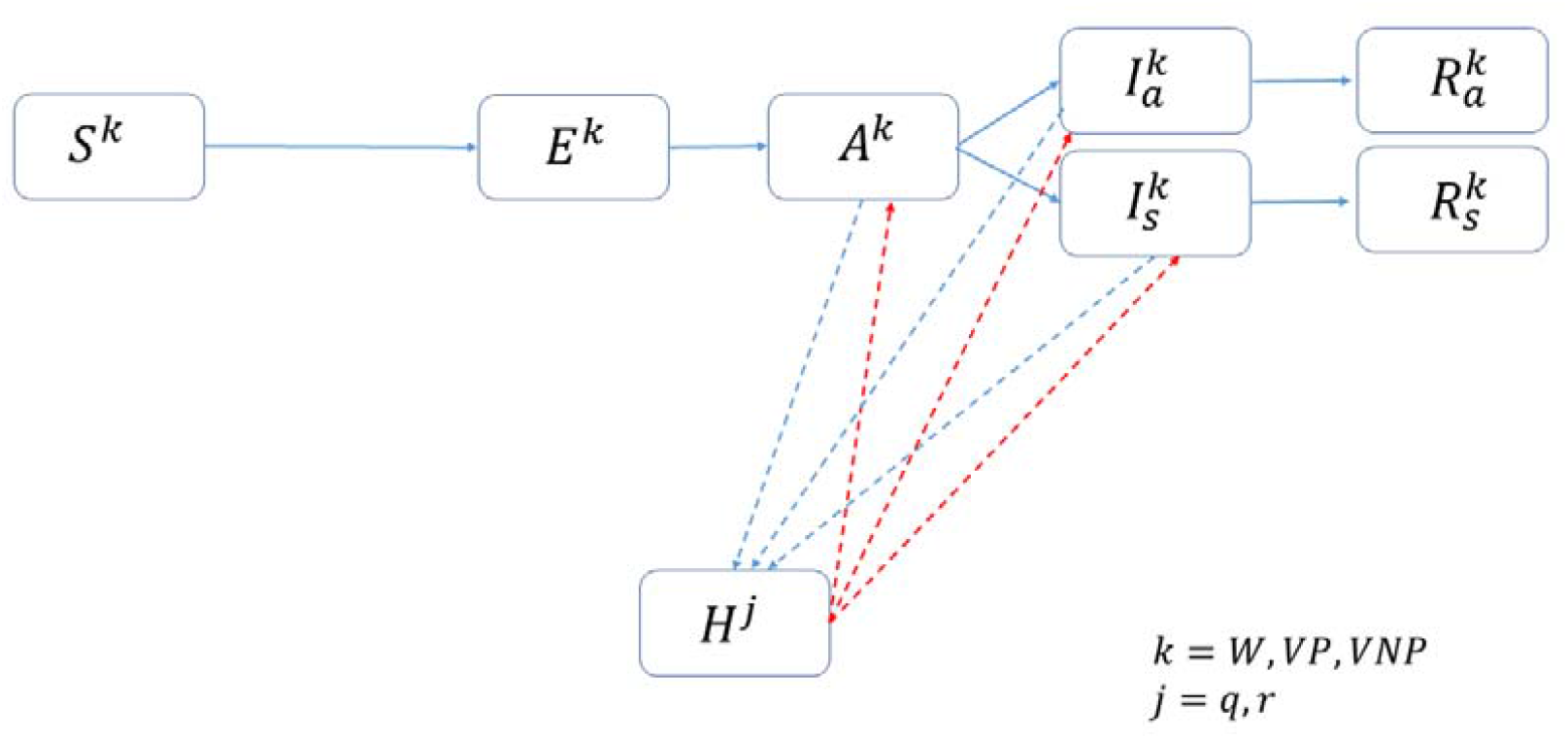
Flow diagram. Disease progression in visitors and workers (solid lines), and connection to environmental reservoirs (dashed lines) is shown. k=W, V$, VNP, j=q, r indicating queues and rides.

Briefly, we consider a multi-patch infectious disease modelling framework with *n* patches. Within each patch *i*, there are worker and visitor populations that are further delineated by disease status: susceptible 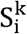, exposed 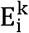, pre-symptomatic infectious 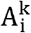, asymptomatic infectious 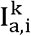, symptomatic infectious 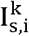 and recovered 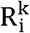 (from infection, or vaccinated), where k denotes the worker W and visitor V populations, and the visitor population is further split into PPE compliant VP and non-compliant VNP groups. We assume that all workers are PPE compliant as this would be part of their job duties. Within each patch, the model also includes an environmental reservoir H, and the patch is further divided into queue *q* and ride *r* locations where both direct (human-human) and indirect (environment-human) transmission can occur.

When individuals present to the park gate, they are first assigned to the worker W, visitor PPE compliant VP, and visitor PPE non-compliant VNP populations. The individual is then assigned a disease status, which depends on the disease prevalence in the surrounding community, including that of the visiting tourist population. If the individual is able to enter the park, that is, they are not showing symptoms and pass any diagnostic test at the park gate, the individual is assigned to a home patch *i* within the park, with a certain probability. Depending on the home patch and whether they are a worker or a visitor, the individual is assigned a proportion of time spent in each of the *n* patches.

The mathematical models track human-human and environment-human pathogen transmission, and pathogen shedding into the environment from infected individuals. It thus tracks the changing infection/disease landscape of all visitors, workers, and environmental reservoirs in the park. Given that some individuals may progress to symptomatic disease, we assume that these individuals will quickly exit the park, minimizing transmission capability. We also assume that the rides and queues will be periodically cleaned throughout the day. When this occurs, we assume that the pathogen load in these environmental reservoirs is set to zero. Finally, we assume that the park closes at the end of day and is cleaned so that the there is no pathogen load the next morning at opening time.

For the ease of mathematical analyses, we consider a variant of the full model with two patches. Given the paucity of data on attraction waiting times in theme-parks, we used published data from [6]. Using this data, we picked the most and least popular attractions, assuming that the ride time in the more popular attraction is slightly longer than that of the least popular one. The waiting times in [6] were extrapolated beyond the reported theme-park operating hours as most of them are open to the general public from 9 am to 9 pm in North America. For simplicity, it is assumed that workers do not switch patches throughout the day and as such remain in their home patch for the entire model duration.

### Basic reproduction number

The transmission of an infection is strictly related to the basic reproduction number, R_0_; the number of secondary cases that an infected individual generates within his/her infectiousness period in a totally susceptible population, without any public health controls. Reported estimates of COVID-19 range from 2-5 in the literature [1, 23]. We explored values of П^NP^ and *κ* that produce similar estimates given different contact rates between workers, visitors, and workers and visitors, c^W^, c^V^, and c^WV^, respectively (5 or 10 contacts are assumed, on average). Parameters П^NP^ and *κ* so that base values of these parameters can be determined given the acceptable range of R_0_ reported in the literature. These values are also free parameters that can be used to inform public health policies within the park.

The *Next Generation Matrix* method [13, 18, 36, 14] is employed to derive the basic reproduction number. Given the complexity of the model, an analytical expression of R_0_ is not feasible, hence it is evaluated numerically

### Sensitivity Analysis

A probabilistic sensitivity analysis using Latin Hypercube Sampling/ Partial Rank Correlation Coefficient (LHS/PRCC) method [25, 24] was undertaken to explore uncertainty in specific model parameters: effectiveness of PPE in halting human-human transmission s_h_; effectiveness of PPE in halting transmission from the environment *f*; proportion of visitors wearing PPE and their effectiveness ν*p*; number of infected individuals entering the park λ*ia*; shedding rate of pathogen into the environment κ; contact rates between workers and visitors c^v^,c^w^,c^wv^; probability of generating new infections in the park between unprotected individuals, and to quantify the effect of these uncertainties on the number of new infections in the park. These are further quantified under different cleaning policies: cleaning once a day before opening, and cleaning multiple times throughout the day. For each scenario, we generate 3000 samples, and the monotonic relationship between model parameters and outputs are verified. Model parameters are varied using uniform distributions. The upper and lower bound values are shown in Table S1.

PRCC values are reported from the LHS/PRCC analysis. These values lie within the range -1 to 1. The sign of the PRCC value provides a measure of the nature of the linear association (correlation) between the parameter and the model outcome being measured. The magnitude of the PRCC value then provides a measure of the strength of this linear association. A relation between model output and a given parameter value is considered significant if the magnitude of the PRCC value is greater than 0.5 [25, 24].

## Results

### Basic reproduction number

Fig. 2 displays the contour plots of R_0_ given parameter space Π^NP^− κ, assuming average contacts between visitors to be 5 or 10, between workers to be 2 and between workers and visitors to be 4. We note that *f=sh= 0* here, as the basic reproduction number is estimated when public health control mechanisms are not in place. Intuitively, as κ and Π^NP^ increase, R_0_ also increases. We also observe that to obtain R_0_ values between 2-5 [1, 23], κ and Π^NP^ must take values in the region bounded by the bold lines (bottom left corner). Herewithin, we take values of κ and Π^NP^ in these ranges.

**Fig. 2.**
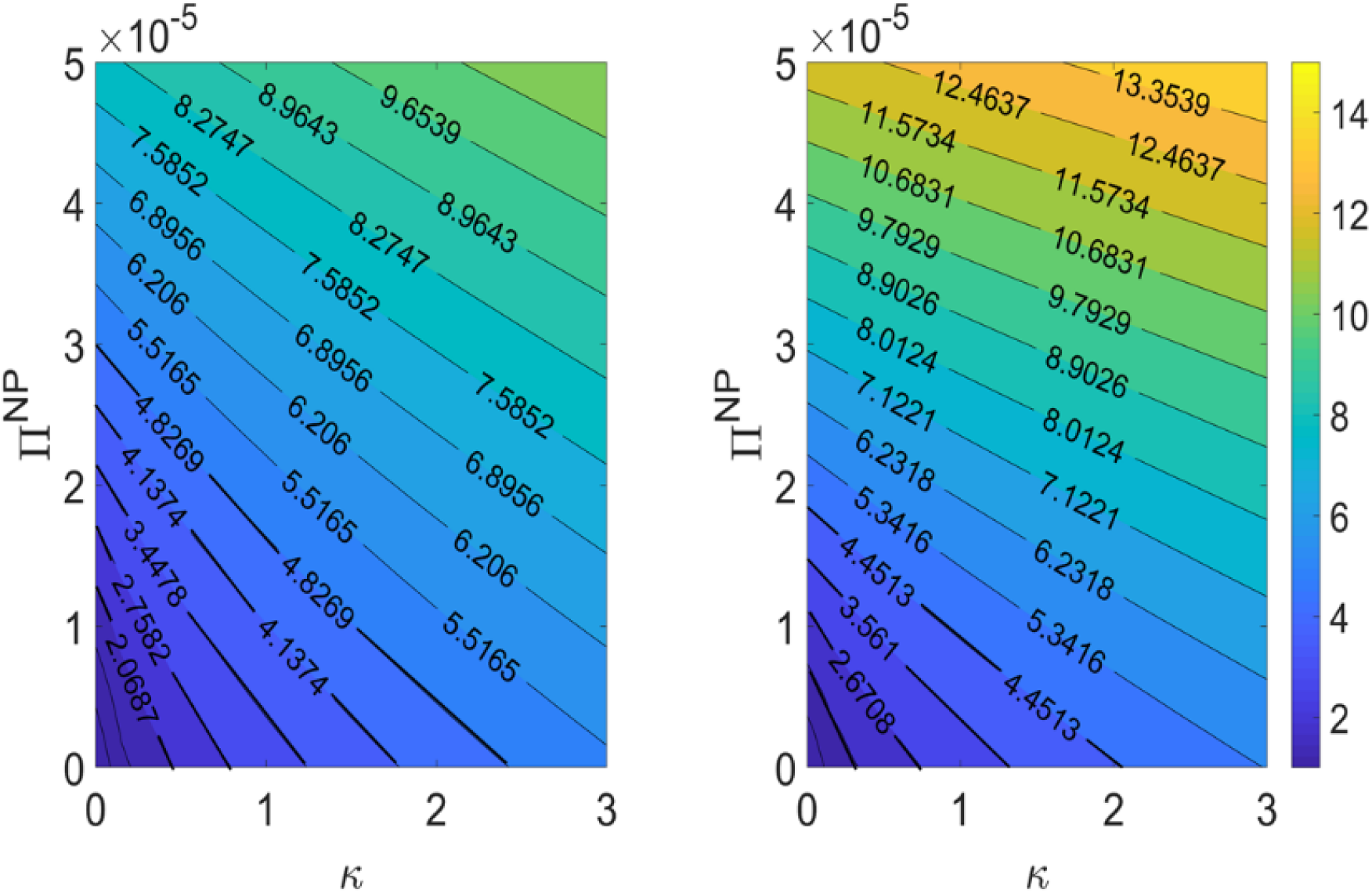
Contour-plots ofR0Contour-plots of the basic reproduction number in Π^NP^−κ parameter space given contacts of (a) 5 or (b) 10 between visitors, 2between workers, and 4 between workers and visitors. Here, f= sh = 0

### Numerical Simulations

In this section, we compare the number of secondary infections A while varying the proportion of infected visitors or workers entering the park *iap*, the proportion of visitors wearing PPE (*vp*), the effectiveness of the PPE (f,*sh*). We also compare model dynamics when cleaning every 2, 3, 4, and 6 hours versus once a day.

Figure3 plots the incidence of new A infections over the course of the opening hours of the theme-park for one such example. Here, f= 0.1,sh= 0.1,ip= 0.05,vp= 0.1 (left panels), 0.5 (middle panels), 0.9 (right panels), the theme-park is cleaned every 2(black lines), 3 (red lines), 4 (magenta lines), or 6 (yellow lines) hours, or when it is cleaned only once a day. Panels (a)-(c) show outcomes when infected individuals are seeded into the visitor population that is not wearing PPE, the visitor population that is wearing PPE, and when infected individuals are distributed between these two classes and the worker population. This figure shows that increased cleaning rates greatly reduce the number of secondary infections A in the park. We also observe reductions in the number of new infected individuals, A, when PPE compliance increases (from left to right). We, however, observe increased cleaning rates have the greatest effect. The costs of cleaning versus provision and enforcement of PPE compliance must therefore be considered by all theme-parks.

We note that the length of the theme-park opening period is less than the average time that it takes for an A to progress to Ia or Is. Additionally, PPE compliance and park cleaning will not have an effect on disease progression. We therefore do not show results concerning these populations here. The effects of PPE and cleaning on transmission of the virus to produce secondary infections is captured in Fig. 3.

**Fig. 3.**
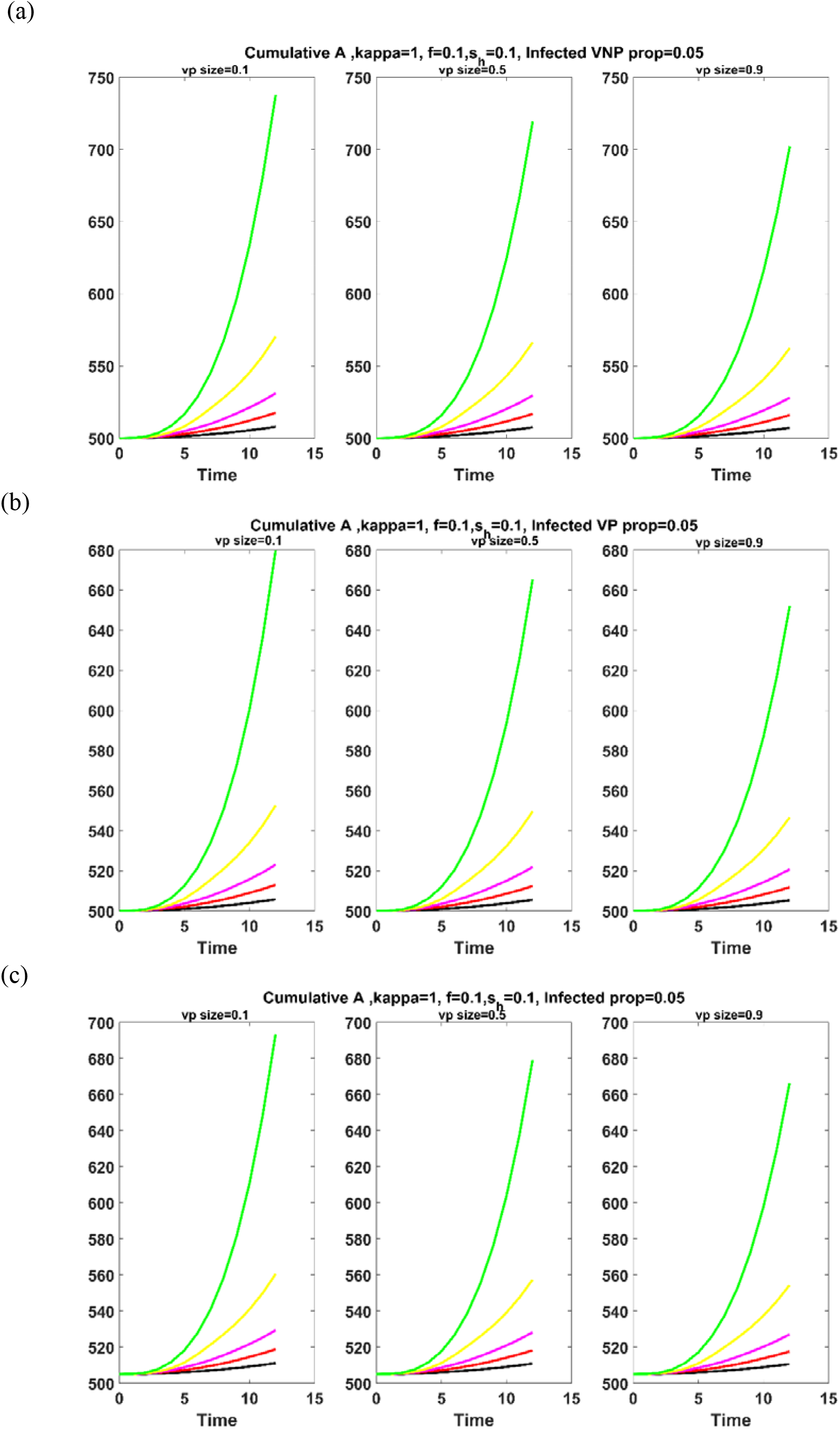
Dynamic of A classes for f= 0.1,sh= 0.1,vpsize= 0.1, 0.5, 0.9 and i_ap_= 0.05 when infected individuals are seeded in the (a) visitors with no PPEs(b) visitors with PPEs, (c) visitors in both PPE classes and a small fraction of workers. The theme-park is cleaned every 2, 3, 4, or 6 hours, or once a day (black, red, magenta, yellow, green, respectively).

### Sensitivity Analysis

In Fig. 4b we plot the results of a sensitivity analysis on the basic and control reproduction numbers, R_0_ and R_C_. This figure shows that R_0_ is significantly affected by the virus shedding rate κ only, and that R_C_ is significantly affected by the virus shedding rate and the effectiveness of PPE against human-environment transmission. These results highlight the importance of reducing the viral load in the environment.

**Fig. 4.**
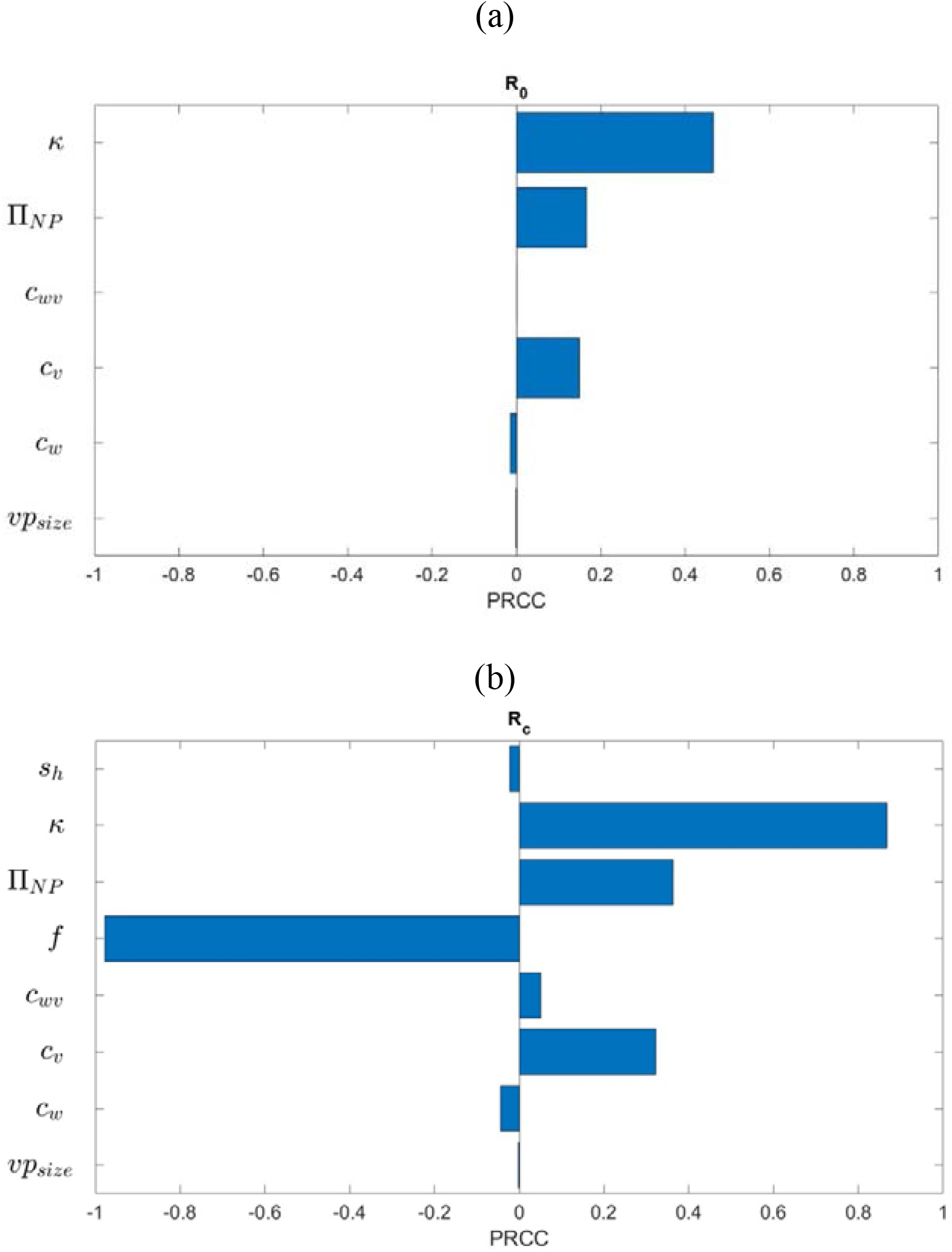
PRCC plots on (a) basic reproduction number (b) control reproduction number

We also studied the impact of cleaning on the cumulative number of new asymptomatic infections, A, over the course of the park day. This is considered under low, medium and high levels of infected individuals entering the park, i_ap_= 0.001−0.05, 0.05−0.1, and 0.1−0.5. Figure 5 shows that when cleaning occurs once a day(left column), reductions in the virus shedding rate κ and the proportion of visitors entering the park i_ap_, and increases in the effectiveness of the PPE in human-environment transmission *f* as well as the proportion of visitors wearing PPE properly vp_size_ are needed to significantly reduce the number of new A infections in the park. When the park is cleaned multiple times a day (right column), to significantly affect the number of new A infections produced in the park, reductions in κ and i_ap_ are still important, as well as an increase inf. However, variations in vp_size_ do not significantly affect the outcome anymore, and instead, the rate of cleaning cl_time_ becomes important. Here, we see that reductions in the number of hours between cleanings are needed to significantly reduce the number of A infections produced over the course of one park day. Finally, we remark that cl_time_ increases in significance as the proportion of infected visitors entering the park increases.

**Fig. 5.**
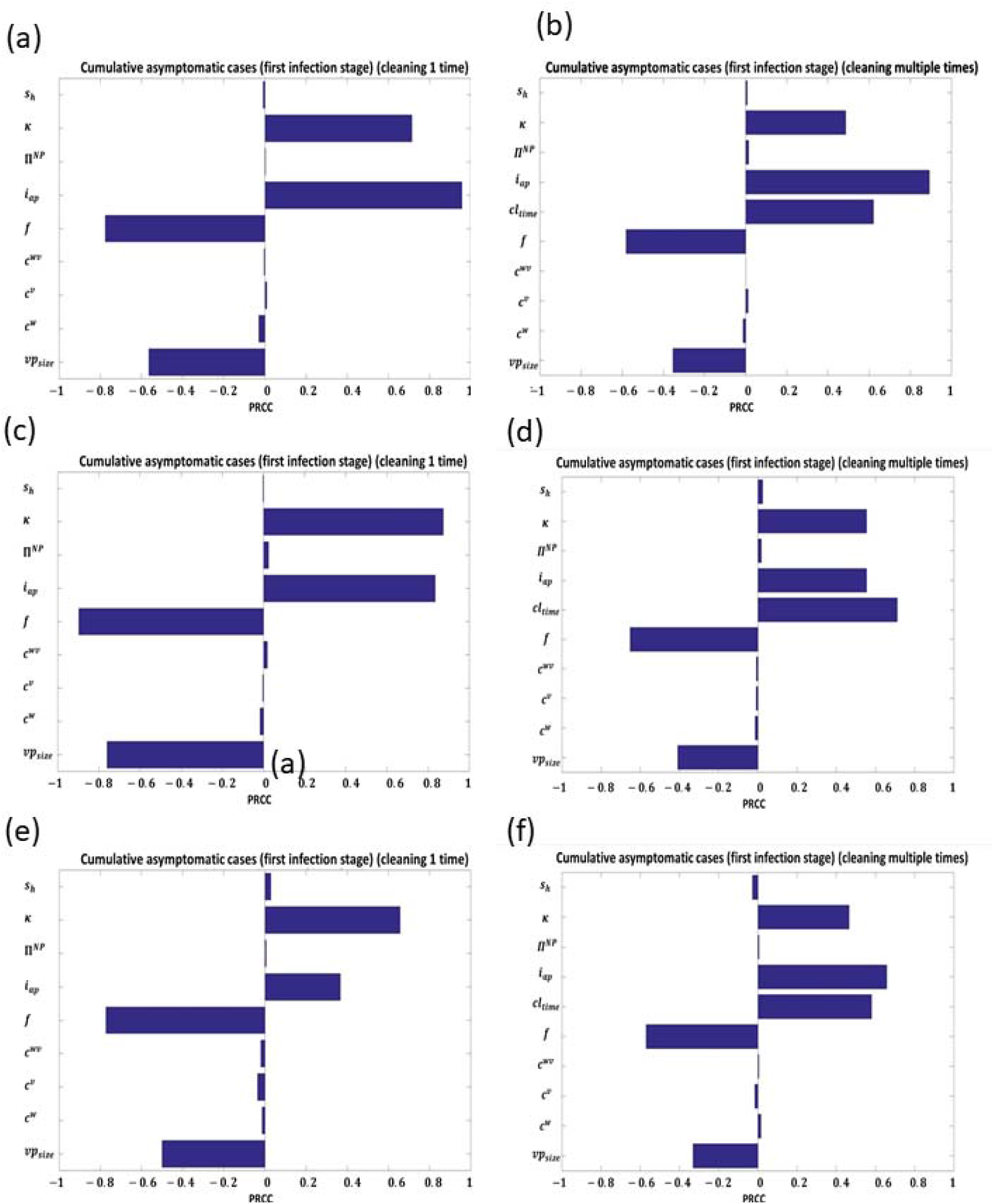
PRCC plots on the number of cumulative asymptomatic A infections produced over the course of one park day when (left panels) cleaning is once a day, (right panels) cleaning occurs multiple times a day, and when the proportion of infected visitors entering the park is (a-b) ip= 0.001−0.05, (c-d) 0.05-0.1, and (e-f) 0.1-0.5.

Given the increase in spread of different variants of concern (VOC) Fall 2020 [2, 42, 10, 40, 20, 12, 28, 41, 12, 28, 41], we have extended our model to include a 1.5 times increase in transmissibility of the virus from infected individuals to other visitors and workers. Fig. 6 plots the results of the sensitivity analysis for this case, again, under low, medium and high levels of infected individuals entering the park (i_ap_= 0.001−0.05, 0.05−0.1, and 0.1−0.5), considering cleaning once a day (left column), or multiple times a day (right column). Comparing to Fig. 5 we see that the sensitivity analysis is not affected by the increase in transmissibility when the incoming rate of infected is low (top row). When the incoming rate of infecteds is increased to 0.05−0.1, we again observe similar results between Figures 5 and 6 given cleaning once a day (panel (c)). However, considering cleaning multiple times a day (panel (d)), there is a change in the order of significance between parameters κ, i_ap_ and cl_time_. These show that the cleaning frequency and environmental shedding becomes more important when transmissibility is increased.

**Fig 6.**
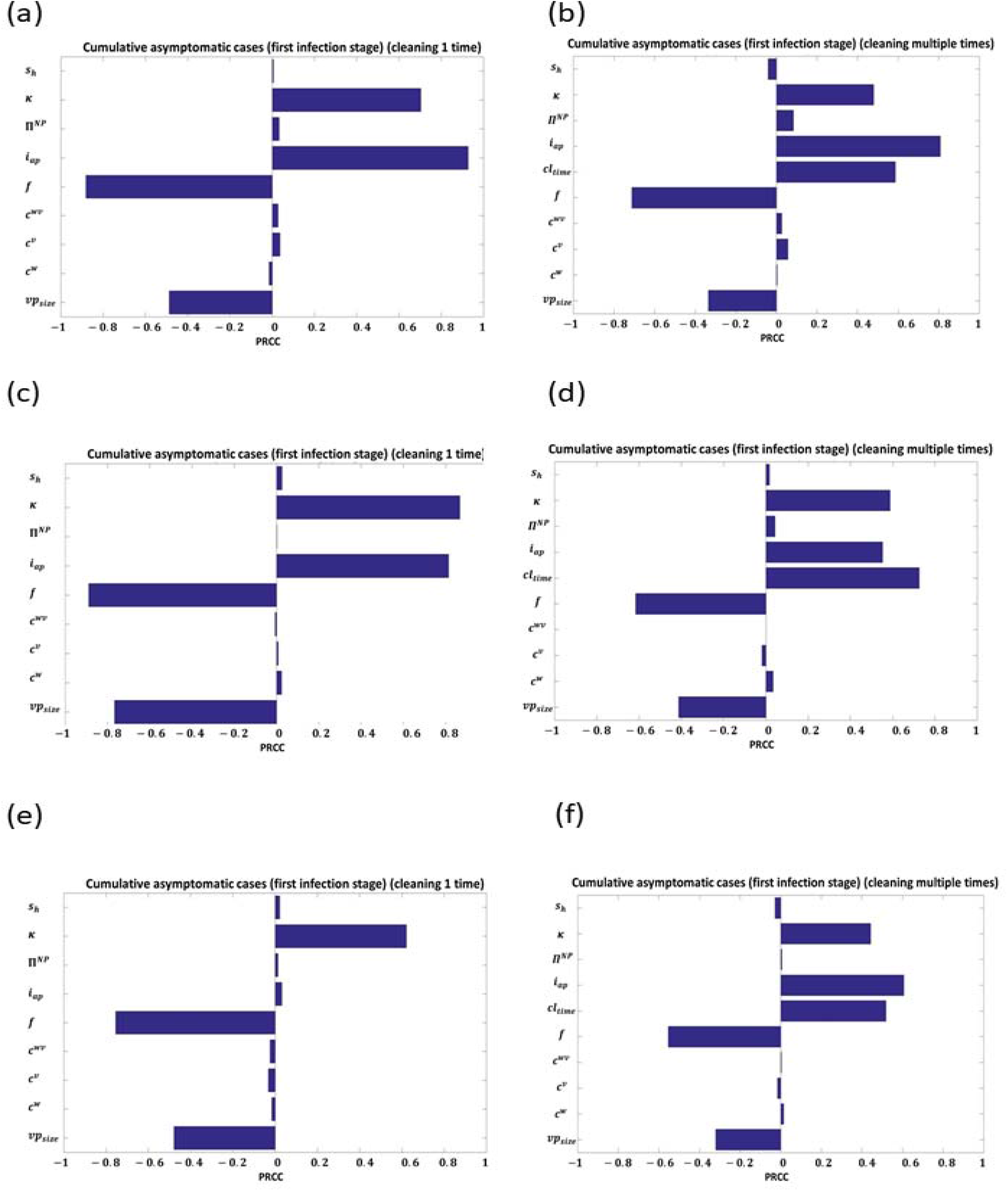
PRCC plots on the number of cumulative asymptomatic A infections produced over the course of one park day when (left panels) cleaning is once a day, (right panels) cleaning occurs multiple times a day, when the proportion of infected visitors entering the park is (a-b) iap= 0.001−0.05, (c-d) 0.05-0.1, and (e-f) 0.1-0.5 and transmission and viral shedding are increased by 50%

## Conclusion

As the world emerges out of lock-down and strict public health intervention measures put in place to contain the COVID-19 pandemic, policy makers around the world are looking for optimal mitigation strategies to limit and control the emergence of new infections in public spaces. theme-parks are starting to open under such conditions. Cleaning of the rides and queues (and other areas of the park),symptom testing at the gate, and mask wearing and hand-washing within the park represent mitigation strategies that a theme-park can adopt to decrease the risk of transmission of the virus. In this study, we have quantified the effects of these mitigation strategies for one theme-park day using a multi-patch model simulating the transmission dynamics of COVID-19 considering both transmission between workers and visitors, and workers and visitors with the environment. The model simulates the interaction between visitors and workers in a theme-park setting while exploring a range of intervention scenarios to reduce disease transmission.

Our results show that reducing virus shedding to the environment κ, increasing the efficacy of PPE in protecting workers and humans from contracting the virus from the environment *f*, increased testing efficacy at the gate so as to decrease the proportion of infected visitors entering the park iap, and increasing the proportion of visitors wearing PPE in the park can all significantly reduce transmission of COVID-19 in a theme-park. We, however, also find that increases in park cleaning rates significantly affect this outcome, and that, as the proportion of visitors entering the park iap increases, the cleaning rate becomes the most important control mechanism for COVID-19 spread. We note, however, that when i_ap_ is low, theme-parks can adopt the strategy of cleaning once a day if they ensure adequate use of PPE (face covering and hand sanitization). We note that as knowledge of waning immunity from vaccination and infections is currently [15] lacking, the wearing of PPE in the park should remain a top priority.

An interesting outcome of the modelling study is that reductions in number of contacts between workers and visitors was not found to be significant in reducing the number of new infections A in the park. This outcome, however, can been explained when considering specific characteristics of direct human-human and indirect environment-human transmission in the park. During a visit to the park, individuals directly contact individuals over very short periods of time during their short visit to the park. Given that the basic reproduction number R_0_ of COVID-19 lies in the interval 2−5 [1, 23], over the lengthy infectious period of COVID-19, the probability of transmitting the virus between humans is small. The shedding rate of the virus to the environment thus lengthens the ’effective infectious period’ of an infected individual, increasing the probability of infection of susceptible. Thus, cleaning rates and mitigation of human-environment contact increases in importance.

The outcomes of this study can guide theme-parks in developing public health mitigation strategies in the park. In the current study we have not considered the costs of implementing COVID-19 testing, park cleaning, and any provision of PPE to park workers and visitors, so we cannot comment on optimal policies that minimize transmission while minimizing cost to the park. This is a course for future work. The current study employs a system of ordinary differential equations. This type of model structure includes broad-sweeping assumptions on the mixing of individuals in the park that ignore spatial aspects of movement, and therefore, may not accurately reflect actual worker and visitor behaviour in the park. Additionally, the existing model ignores different transmission probabilities between indoor and outdoor venues. In current work we are implementing agent-based models using specific park structures with pathways, queues and rides that better represent theme-park spatial structure, indoor and outdoor spaces, and movement of the workers and visitors.

## Data Availability

The source of data is cited in the manuscript.

## Acknowledgments

This work is supported by NSERC (JMH, JW, AA), an NSERC-Sanofi Industrial Research Chair (JW, JMH, AA), and the York University Research Chair program (JMH).

## Author Contributions

Study design: EA, SA, AR, JMH. Analysis and simulations: EA, SA, AR, JDK, JMH. Writing and revision of manusript: EA, SA, AR, MAA, MA, CC, MR, CL, JDK, JMH. Advising: JW, AA, JDK, JMH. Supervision: JMH.

## SUPPLEMETARY MATERIAL

### A. Model Description

A detailed flow diagram of the model is provided in Figure S1. Model parameters are listed in Table S1. Visitors and workers present to the park at the park gate. At the gate, they will be monitored for symptoms. If they are clear of symptoms, they will be allowed to enter the park. Thus, only susceptible, pre-symptomatic, asymptomatic infectious, and recovered individuals will be allowed to enter. We assume that *λ*_P,i_ determines the fraction of individuals that enter the park in susceptible, the asymptomatic and pre-symptomatic stages of infection.

We assume that the park is split into *n* patches. Once a worker or visitor has entered the park, they are assigned a home patch. Visitors are also assigned to PPE compliant and non-compliant subgroups, VP and VNP, with probabilities v_p_ and 1-v_p_ respectively. This assignment is random and is varied in our simulations. We assume that workers are PPE compliant as per employer guidelines.

The assignment of the home patch ultimately determines the fraction of time y_ij_^k^ that an individual will spend in each of the *n* patches during their time in the park, where *y = w, v, v*^*Is*^, *i, j = 1*,…,*n, k=W, VP, VNP*, and 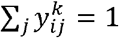 for any combination *of i=1*,…,*n* and *k=W, VP, VNP*. The distribution of the home patch assignments can be provided by the theme-park. Currently, we assume a distribution 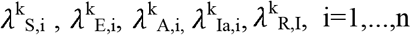 for each k = W, V, where V is the total visitor population (VP+VNP).

Susceptible individuals, S, can be infected by pre-symptomatic, asymptomatic, and symptomatic infectious workers and visitors from any patch (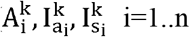, k=W, VP, VNP) in the park, or indirectly through a contaminated environment in a queue or on a ride (H^q^,H^r^)Once infected, an individual moves to the exposed class E and remains there for 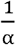 days. Subsequently, an exposed individual moves to the pre-symptomatic infectious class 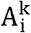 and remains there for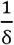 days. Following disease progression, a COVID-19 infected individual can either become symptomatically infected, 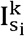 with probability q, or stay asymptomatic, 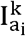, with a probability 1-q. Upon manifestation of symptoms, we assume that visitors and workers will leave the park within 30 minutes 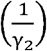

Transmission of the pathogen between humans is modelled using contact rates between workers c^W^, visitors c^V^, and workers and visitors c^WV^, and transmission probabilities between PPE compliant individuals П^*P*^, PPE non-compliant individuals П^*NP*^ and between individuals that are from the PPE compliant and non-compliant groups П^*PNP*^. We assume that П^*P*^ and П^*PNP*^ are related to П^*NP*^ and the effectiveness of PPE in preventing human-human transmission s_h_. Transmission to humans from the environment is modelled using transmission rates in the queues 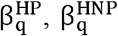, and rides 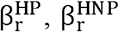 given individuals that are PPE compliant (HP) or non-compliant (HNP). Additionally, we assume that infectious individuals can shed pathogen into the queue and ride environments, with rate *κ*, and we assume that PPE have *f* effectiveness in reducing this shedding.

### B. Model Equations

For a given population *k = W, VP, VN* and a given patch *i=1*,..,*n*, we have the following differential equations:

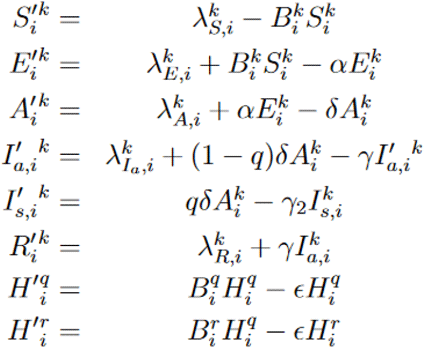

where

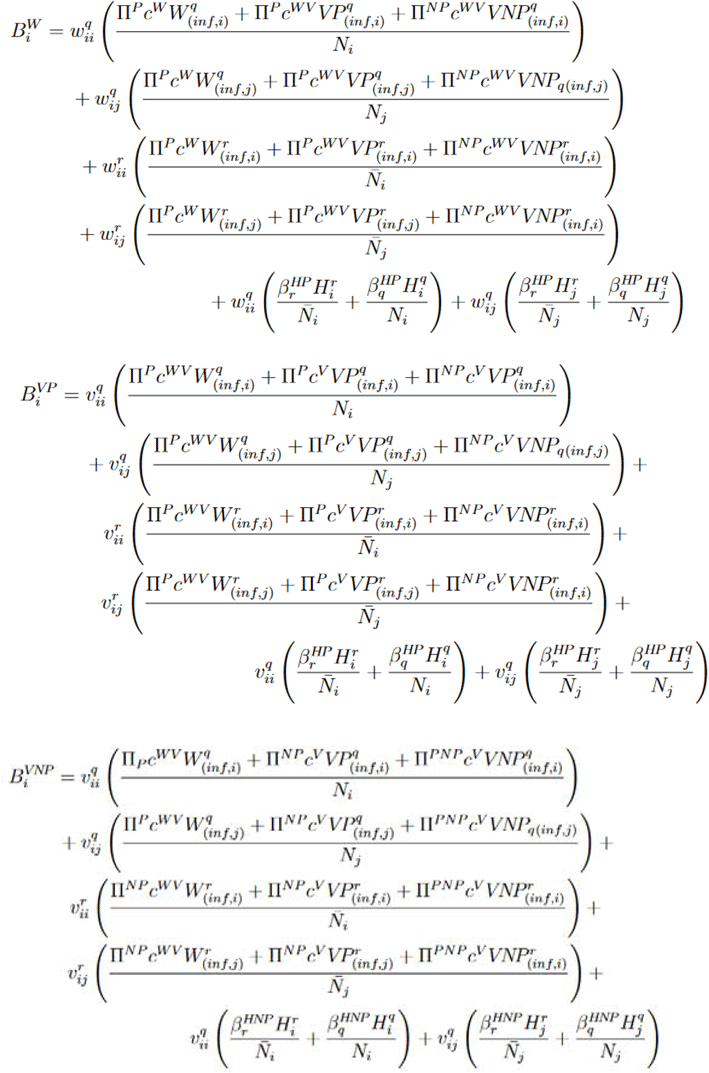

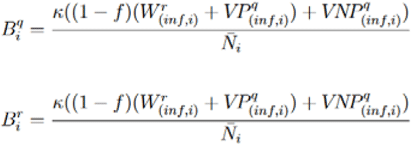

Here infection in queue for workers, visitors with PPE (VP) and visitors without PPE (VNP) where i,j∈{1,2,…,n}

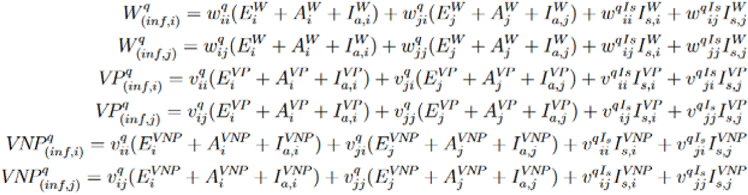

And infection on Rides for workers, visitors with PPE (VP) and visitors without PPE (VNP) where when i,j∈{1,2,…,n}, are given as follows:

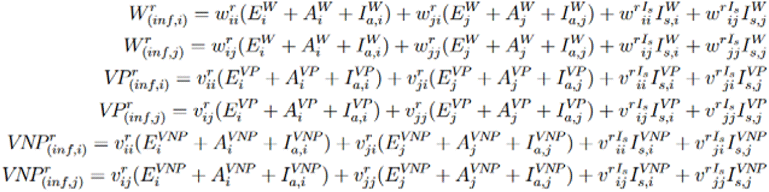

**Table S1:**
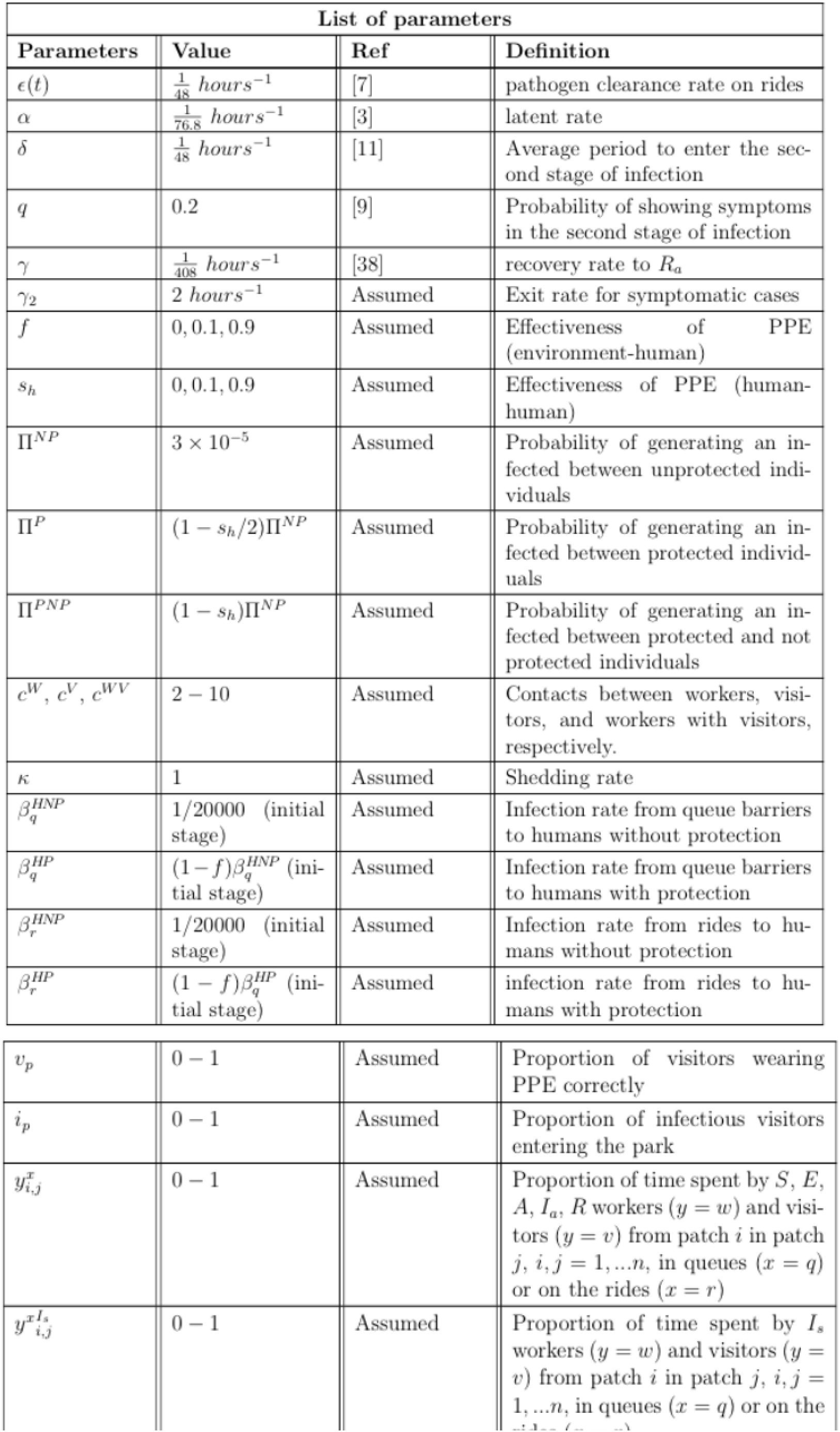
Table of Parameters.

| List of parameters |  |  |  |
| --- | --- | --- | --- |
| Parameters | Value | Ref | Definition |
| $\epsilon(t)$ | $\frac{1}{48} \text{ hours}^{-1}$ | [7] | pathogen clearance rate on rides |
| $\alpha$ | $\frac{1}{76.8} \text{ hours}^{-1}$ | [3] | latent rate |
| $\delta$ | $\frac{1}{48} \text{ hours}^{-1}$ | [11] | Average period to enter the second stage of infection |
| $q$ | 0.2 | [9] | Probability of showing symptoms in the second stage of infection |
| $\gamma$ | $\frac{1}{408} \text{ hours}^{-1}$ | [38] | recovery rate to $R_a$ |
| $\gamma_2$ | $2 \text{ hours}^{-1}$ | Assumed | Exit rate for symptomatic cases |
| $f$ | 0, 0.1, 0.9 | Assumed | Effectiveness of PPE (environment-human) |
| $s_h$ | 0, 0.1, 0.9 | Assumed | Effectiveness of PPE (human-human) |
| $\Pi^{NP}$ | $3 \times 10^{-5}$ | Assumed | Probability of generating an infected between unprotected individuals |
| $\Pi^P$ | $(1 - s_h/2)\Pi^{NP}$ | Assumed | Probability of generating an infected between protected individuals |
| $\Pi^{PNP}$ | $(1 - s_h)\Pi^{NP}$ | Assumed | Probability of generating an infected between protected and not protected individuals |
| $c^W, c^V, c^{WV}$ | 2 – 10 | Assumed | Contacts between workers, visitors, and workers with visitors, respectively. |
| $\kappa$ | 1 | Assumed | Shedding rate |
| $\beta_q^{HNP}$ | 1/20000 (initial stage) | Assumed | Infection rate from queue barriers to humans without protection |
| $\beta_q^{HP}$ | $(1 - f)\beta_q^{HNP}$ (initial stage) | Assumed | Infection rate from queue barriers to humans with protection |
| $\beta_r^{HNP}$ | 1/20000 (initial stage) | Assumed | Infection rate from rides to humans without protection |
| $\beta_r^{HP}$ | $(1 - f)\beta_q^{HP}$ (initial stage) | Assumed | infection rate from rides to humans with protection |
| $v_p$ | 0 – 1 | Assumed | Proportion of visitors wearing PPE correctly |
| $i_p$ | 0 – 1 | Assumed | Proportion of infectious visitors entering the park |
| $y_{i,j}^x$ | 0 – 1 | Assumed | Proportion of time spent by $S, E, A, I_a, R$ workers ($y = w$) and visitors ($y = v$) from patch $i$ in patch $j, i, j = 1, \dots, n$, in queues ($x = q$) or on the rides ($x = r$) |
| $y_{i,j}^{xI_s}$ | 0 – 1 | Assumed | Proportion of time spent by $I_s$ workers ($y = w$) and visitors ($y = v$) from patch $i$ in patch $j, i, j = 1, \dots, n$, in queues ($x = q$) or on the rides ($x = r$) |

**Figure S1:**
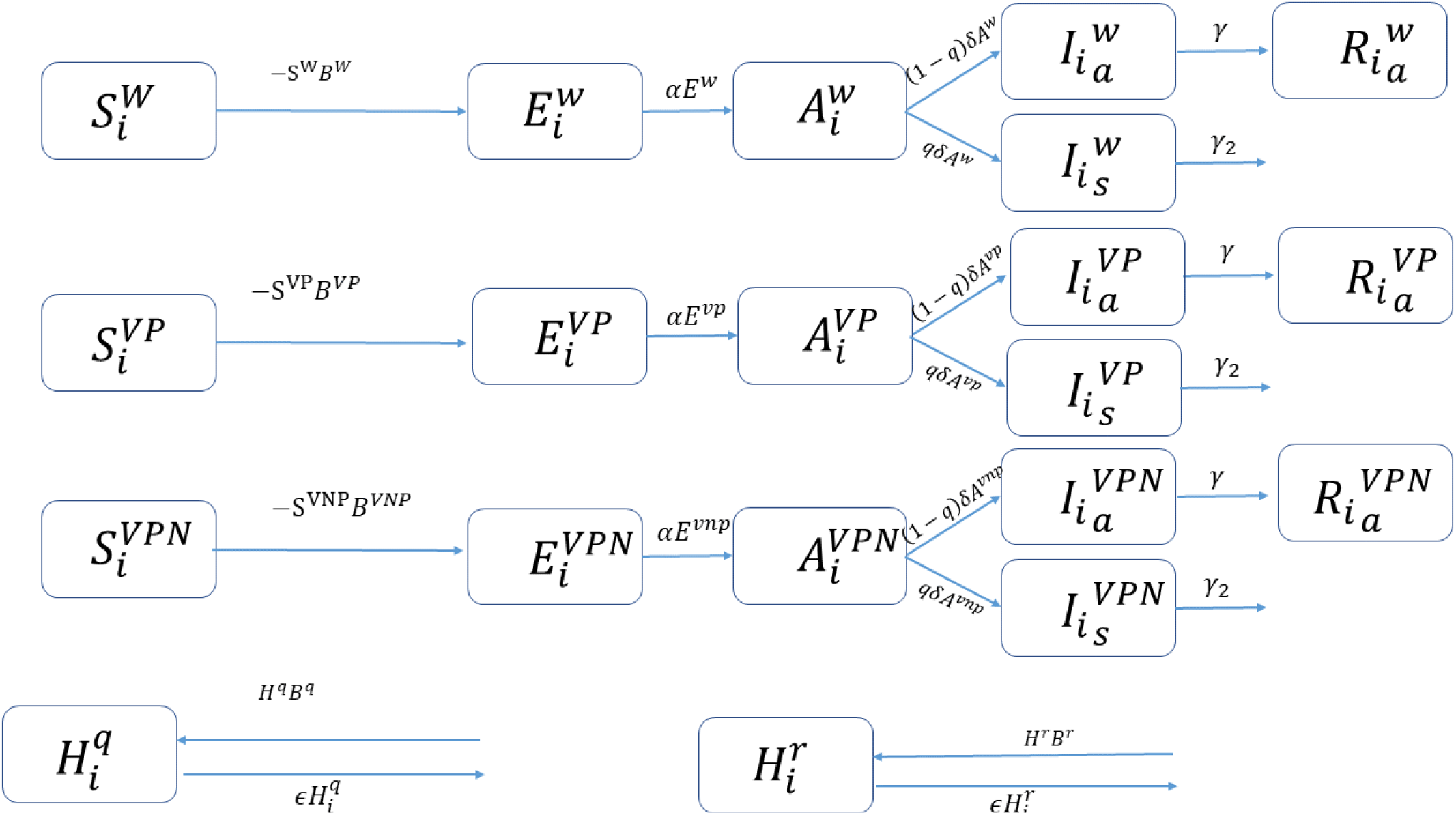
Flow diagram of Systems S1, in patch i

